# Middle-Aged and Older Adults’ Knowledge, Ratings, and Preferences for Receiving Multicomponent Lifestyle-Based Brain Health Interventions

**DOI:** 10.64898/2025.12.02.25341494

**Authors:** Raymond L Ownby, Gesulla Cavanaugh, Shannon Weatherly, Shazia Akhtarullah, Joshua Caballero

**Affiliations:** Department of Psychiatry and Behavioral Medicine, Nova Southeastern University, Fort Lauderdale, Florida USA; Ron and Kathy Assaf College of Nursing, Nova Southeastern University, Fort Lauderdale, Florida USA; Department of Clinical and Administrative Pharmacy, University of Georgia, Athens, Georgia USA

**Keywords:** brain health, cognitive aging, lifestyle interventions, middle-aged adults, older adults, health behavior, dementia prevention, health preferences, survey research, multicomponent interventions, health literacy, preventive health behavior

## Abstract

**Objectives:** Lifestyle behaviors such as physical activity, cognitive engagement, social interaction, diet, sleep, and vascular risk management are increasingly recognized as contributors to cognitive aging and dementia risk. Although many middle-aged and older adults express interest in maintaining brain health, less is known about their detailed knowledge of brain-healthy behaviors or their preferences for receiving multi component brain health interventions. This study examined adults’ ratings of the usefulness of a wide range of lifestyle activities for brain health and their preferred formats for receiving support

**Methods:** A 60-item online survey was administered to compensated volunteers aged 40 years and older through a commercial provider. The questionnaire assessed perceived usefulness of lifestyle-based brain health activities and preferred intervention delivery formats. The analytic sample included 761 respondents. Descriptive statistics were computed for all ratings and differences by age group and gender were tested using MANOVA with post-hoc comparisons adjusted for multiple testing.

**Results:** Participants endorsed many lifestyle activities as helpful for brain health. Mentally stimulating activities, good sleep, stress management, and creative activities received the highest ratings, whereas strength training, meditation, language learning, and computer-based cognitive training were rated lower. Aerobic exercise and mentally stimulating activities were most frequently selected as the single most important activity. Significant effects of age, gender, and their interaction were observed, with younger men and older women generally rating activities more favorably. With respect to desire for services, over half of participants preferred receiving a cognitive assessment, and many favored online education or app-based tools.

**Conclusions:** Middle-aged and older adults recognize a wide range of lifestyle factors as potentially beneficial for brain health and express strong interest in structured support, particularly assessments and digital resources. These findings can inform the design of flexible, multicomponent brain health interventions aligned with adults’ preferences and priorities.

## 1. Introduction

As demographic shifts lead to an increase in aging population [1], there is a growing focus on interventions that can help to sustain cognitive health including memory and higher-order thinking abilities. It is common for adults in their 40s, 50s, and beyond to say they want to “keep their brains sharp” or “avoid dementia,” but they are often unsure what that means in practical terms and have an exaggerated view of their risk for developing dementia [2]. Research over the last decade has shown that lifestyle factors such as physical activity, cognitive engagement, social connection, diet, and management of vascular risk can risk for cognitive aging and dementia [3, 4]. At the same time, messages about “brain health” in the media can be inconsistent or oversimplified. Many adults know that “staying active” is good for the brain [5], but they may not know which behaviors matter most, how much is enough, or how to combine different activities in a realistic way [6, 7].

Studies of older adults’ knowledge of risk and protective factors for brain health have mixed results. In several surveys and qualitative studies, older adults endorsed lifestyle factors as important for cognitive health, including physical activity, mental stimulation, social interaction, and diet [5, 7, 8]. Many participants in these studies also recognized that dementia risk is not entirely fixed and believed that at least some risk is modifiable [7, 9]. At the same time, detailed knowledge of specific risk factors and recommended behaviors is often limited. Older adults commonly note that increasing aging and genetics are key risks for dementia but are less aware of the roles of hypertension, obesity, smoking, and other cardiovascular factors [10, 11]. Knowledge appears to vary by education and socioeconomic status, with some studies reporting lower awareness among those with less education or lower income [8, 11]. Cultural variations shape conceptualizations of brain health, with certain groups prioritizing spiritual or community-oriented approaches, while others predominantly depend on interpersonal networks and informal communication channels for health information [12].

Collectively, these studies suggest that middle-aged and older adults recognize the importance of lifestyle factors for brain health and can identify several activities perceived as beneficial. They also indicate persistent gaps and inconsistencies in knowledge, particularly around vascular risk factors, the specific characteristics of effective activities (for example, how often and at what intensity to exercise), and the role of less visible factors like sleep and mood. Importantly, most of this work has focused on what people know, what they believe about risk and prevention, or how motivated they say they are to change. Much less attention has been paid to how adults would like to receive help in making their lifestyles more brain healthy. Only a few studies have touched on preferences for different kinds of interventions or support (e.g., education vs. counseling vs. technology-based programs), and these have typically not examined preferences across a broad range of multicomponent brain health activities [13].

Although prior studies, including ours [14], have outlined what middle-aged and older adults know about brain health and which behaviors they consider important, there has been little rigorous examination of their preferences regarding intervention design and implementation formats. What remains largely unstudied is whether adults who are concerned about their brain health prefer self-guided tools versus structured programs, in-person versus remote formats, or individual versus group support, and how they prioritize different activities if they cannot do everything at once. In addition, relatively little is known about what middle-aged and older adults are already doing in their everyday lives to support brain health, beyond broad self-reports of being active.

The present study was designed to address this gap. We surveyed a large sample of middle-aged and older adults to assess their knowledge and beliefs about activities that may support brain health, their current engagement in these activities, and their preferences for receiving multicomponent, lifestyle-based brain health interventions. Participants rated the perceived helpfulness of a range of brain health activities and indicated how they would most like to receive support, for example, through online or in-person education, group programs, mobile apps, or cognitive assessments. We also examined whether ratings and preferences varied by age group and gender. By linking what people know and how they would prefer to receive services, this study aims to inform the design of brain health interventions that are better aligned with the concerns and everyday realities of middle-aged and older adults.

## 2. Materials and Methods

To address this gap in knowledge, a survey was developed to assess middle-aged and older adults’ beliefs about the helpfulness of various brain health activities, including aerobic exercise, healthy diet, depression treatment, and the use of supplements. Activities were drawn from review articles on risk and protective factors for cognitive decline and dementia [4], professional association guidelines [15, 16], and brain health experts [17-20] with the overall goal of assessing a broad array of activities or interventions while still keeping participant response burden at an acceptable level. In addition to rating these activities and to better understand preferences for implementation, participants also selected preferred delivery formats, such as online or in-person programs.

Sample size was estimated using Cochran’s equation for survey research sample size [21] targeting 99% confidence level and 5% margin of error (required n = 664). The target was to obtain stable estimates across key demographic subgroups including age and gender. We also conducted secondary power analyses for planned chi-square tests examining associations between age group, gender, and ratings of activity helpfulness for a range of test degrees of freedom from 1 to 5 (for example, analyzing age group by gender) using routines available in PASS 15 [22]. These analyses showed that a sample size of approximately 700 participants would provide power greater than 0.90 to detect small-to-moderate effect sizes (W = 0.15–0.20) at an alpha level of 0.05.

The survey was distributed to compensated volunteers aged 40 years and older via a commercial survey provider (San Matteo, California, USA: SurveyMonkey Audience). The survey was initially piloted with a target sample size of 100; we collected 160 responses. As the survey had a 22% abandonment rate, we reviewed questions for clarity and to reduce possible confusion. The final questionnaire included 60 questions, asking participants for ratings of usefulness of activities and their preferences for receiving brain health support (see Supplementary Material for the complete questionnaire).

Responses were analyzed to provide a description of participants’ responses and examine differences in ratings by age and gender, as well as overall patterns in knowledge and preferences. Descriptive statistics for items and ratings of the helpfulness of brain health activities were evaluated by age and gender via MANOVA in SPSS 29 (Armonk New York, USA: IBM) with post-hoc tests adjusted for multiple comparisons by Student-Newman-Keuls. Multiple comparisons for between-subjects differences for age and gender and their interaction were Bonferroni corrected.

### 2.1. Human Subjects

This study was completed under a protocol approved by the Institutional Review Board of Nova Southeastern University (protocol number NSU-2024-10). Participants were required to review an online consent form detailing the purpose of the study and asked to provide their consent for participation after its review before responding to the survey.

## 3. Results

The survey received 792 responses, with an abandonment rate of 16%. Responses from individuals younger than 40 years (n = 31) were excluded from the analytic sample. The final sample included 761 individuals (mean age = 52.6 years); 59% were women. Participants were categorized into early middle age (40–50 years, n = 380), late middle age (51–64 years, n = 265), and older adult (≥ 65 years, n = 113) groups. The sample included 644 Whites, 54 Blacks, 44 Asian or Pacific Islander, 10 American Indican or Alaskan Native, and 5 persons with multiple races. Hispanic ethnicity was reported by 99 participants. The sample was highly educated, with 481 reporting a college degree or greater level of education, and 273 reporting less than a college degree.

### 3.1. Ratings of Activities

Participants’ ratings of the usefulness of activities are listed in Table 1. The number responding to each is less than the total number of participants because participants were provided with the option to respond “I don’t know” and “Prefer not to answer.”

**Table 1.** Ratings of Activity Usefulness.

|  | N | Mean | Std. Deviation |
| --- | --- | --- | --- |
| Aerobic exercise | 733 | 3.94 | 1.07 |
| Strength or resistance training | 681 | 3.55 | 1.20 |
| Mentally stimulating activities | 750 | 4.23 | 0.95 |
| Using a computer several times a week | 732 | 3.72 | 1.15 |
| Learning a language or musical instrument | 651 | 3.81 | 1.15 |
| Following a diet rich in fruits and vegetables | 723 | 4.03 | 1.04 |
| Computer-based cognitive training | 655 | 3.71 | 1.13 |
| Getting together once a month or more often with friends or family | 731 | 3.94 | 1.09 |
| Creative activities like painting, music, or writing | 719 | 4.12 | 0.97 |
| Meditating | 691 | 3.72 | 1.13 |
| Getting regular sleep | 742 | 4.27 | 0.98 |
| Reducing alcohol use | 683 | 4.03 | 1.14 |
| Stopping or never smoking | 665 | 4.15 | 1.17 |
| Managing stress | 733 | 4.26 | 0.92 |
| Regular medical checkups | 739 | 4.09 | 1.05 |
| Maintaining a healthy weight | 727 | 3.95 | 1.06 |
| Coping with depression or not being depressed | 711 | 4.07 | 1.08 |
| Getting a hearing test or using a hearing aid if you have trouble hearing | 622 | 3.86 | 1.19 |
| Taking brain health supplements | 651 | 3.64 | 1.23 |

Note: N represents number of valid ratings, as participants could also answer “Don’t know” and “Prefer not to answer”

As illustrated by ratings in Table 1, mentally stimulating activities, getting regular sleep, and managing stress were the highest rated activities, while strength or resistance training, computer use, learning a language or musical instrument, computer-based cognitive training, and meditating were ranked lowest.

Multivariate tests in the MANOVA indicated significant effects of age group (Wilks’ lambda =.81, *F* (38, 878) = 2.45, *p* < 0.001) and the interaction of gender with age (Wilks’ lambda =.15, F (38, 878) = 1.91, *p* < 0.001) on ratings of activity effectiveness. In tests of between-subjects effects, significant differences across age groups were found for rating of strength training (*F* [2] = 6.82, *p* = 0.001), creative activities (*F* [2] = 6.50, *p* = 0.002), meditation (*F* [2] = 9.24, *p* < 0.001), and supplements (*F* [2] = 14.04, *p* < 0.001). In each case, younger individuals rated each of these activities as more useful than older groups (see Figure 1). Between-subjects comparisons for the effects of gender resulted in between-group differences only for hearing (*F* [2] = 5.48, *p* = 0.02), with women rating its usefulness higher than men (4.06 vs 3.76; Bonferroni-adjusted *p* = 0.02).

**Figure 1.**
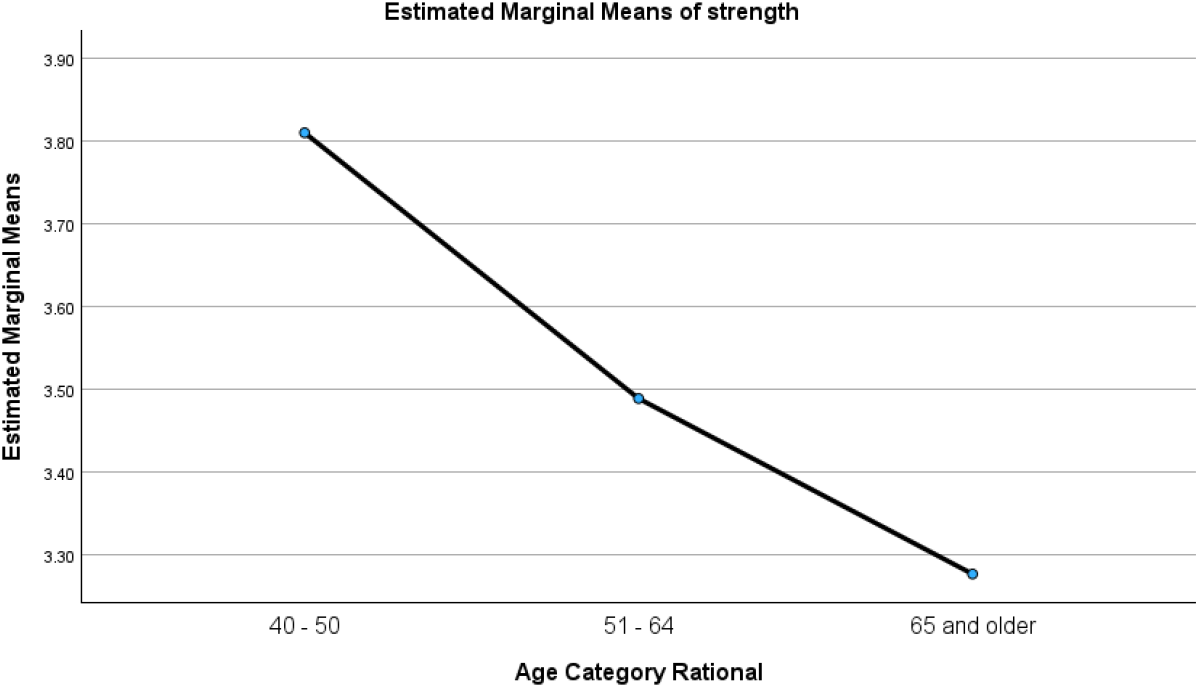
Ratings of Strength Training Over Age Groups

A large number of interaction effects were statistically significant (see Table 2). In general, these effects resulted from younger men rating activities higher than older men, and older women rating activities higher than younger women (see, for example, Figures 2 and 3). Taken together, these findings suggest that age and gender shape perceptions of brain health activity usefulness among middle-aged and older adults. Younger men tended to rate physically demanding activities such as aerobic and strength training more favorably, whereas older women were more likely to endorse preventive and health-monitoring activities.

**Table 2.** Interactions of Age and Gender with Ratings of Usefulness.

|  | Sum of Squares | *df* | Mean Square | *F* | *p* |
| --- | --- | --- | --- | --- | --- |
| Aerobic exercise | 9.78 | 2 | 4.89 | 4.94 | **0.008** |
| Strength or resistance training | 18.02 | 2 | 9.01 | 6.96 | **0.001** |
| Mentally stimulating activities | 3.14 | 2 | 1.57 | 1.85 | 0.158 |
| Using a computer several times a week | 18.00 | 2 | 9.00 | 7.19 | **0.001** |
| Learning a language or musical instrument | 6.01 | 2 | 3.00 | 2.43 | 0.089 |
| Following a diet rich in fruits and vegetables | 5.19 | 2 | 2.59 | 2.70 | 0.069 |
| Computer-based cognitive training | 26.72 | 2 | 13.36 | 10.99 | **< 0.001** |
| Getting together once a month or more often with friends or family | 26.16 | 2 | 13.08 | 12.43 | **< 0.001** |
| Creative activities like painting, music, or writing | 13.15 | 2 | 6.57 | 7.97 | **< 0.001** |
| Meditating | 25.35 | 2 | 12.68 | 11.36 | **< 0.001** |
| Getting regular sleep | 6.05 | 2 | 3.03 | 3.44 | **0.033** |
| Reducing alcohol use | 12.82 | 2 | 6.41 | 5.94 | **0.003** |
| Stopping or never smoking |  | 2 | 1.48 | 1.17 | 0.313 |
| Managing stress | 4.53 | 2 | 2.27 | 2.96 | 0.053 |
| Regular medical checkups | 12.34 | 2 | 6.17 | 6.15 | **0.002** |
| Maintaining a healthy weight | 16.27 | 2 | 8.14 | 7.81 | **< 0.001** |
| Coping with depression or not being depressed | 16.88 | 2 | 8.44 | 7.98 | **< 0.001** |
| Getting a hearing test or using a hearing aid if you have trouble hearing | 20.33 | 2 | 10.16 | 8.05 | **< 0.001** |
| Taking brain health supplements | 34.27 | 2 | 17.13 | 13.81 | **< 0.001** |

**Figure 2.**
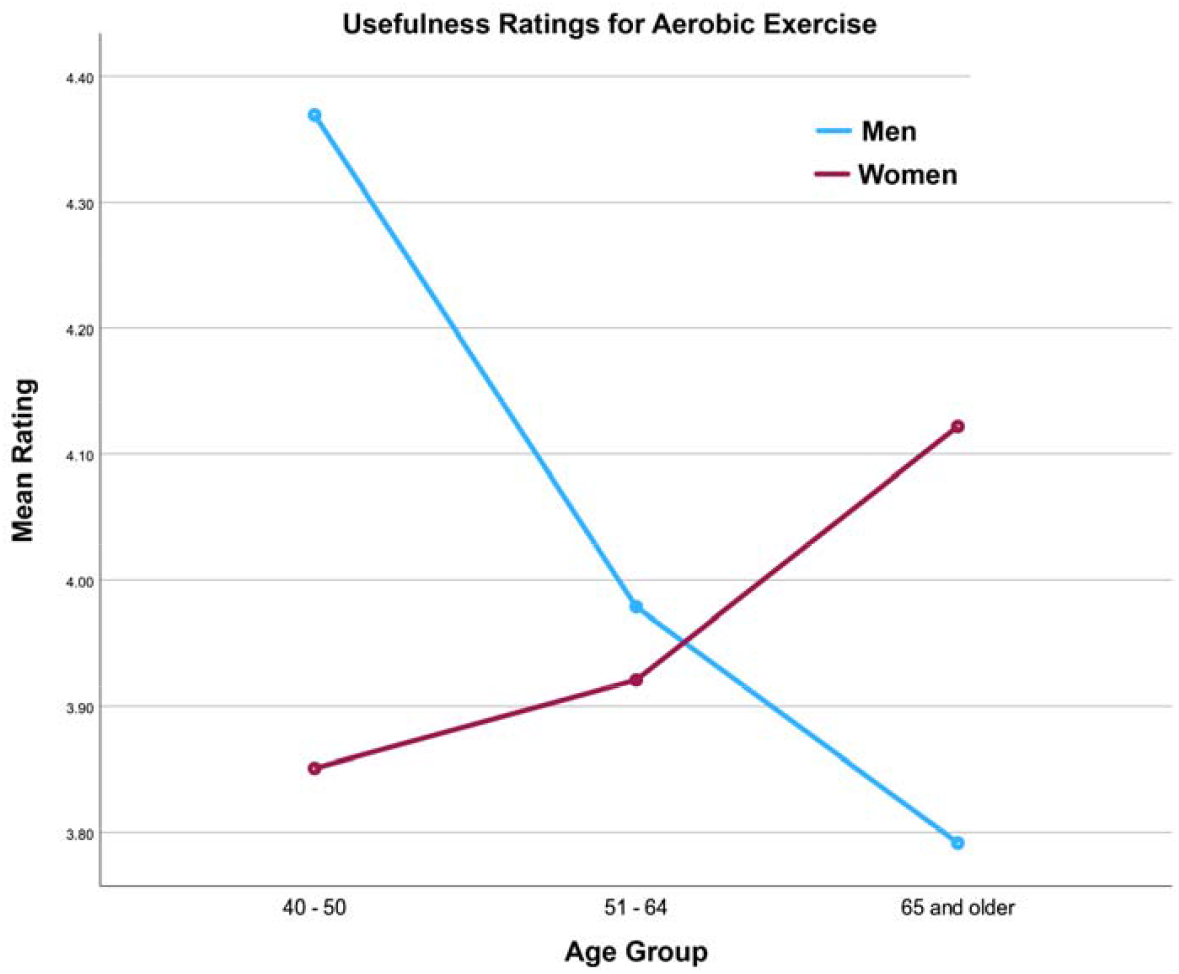
Interaction Between Age and Gender in Ratings of the Usefulness of Aerobic Exercise for Brain Health

**Figure 3.**
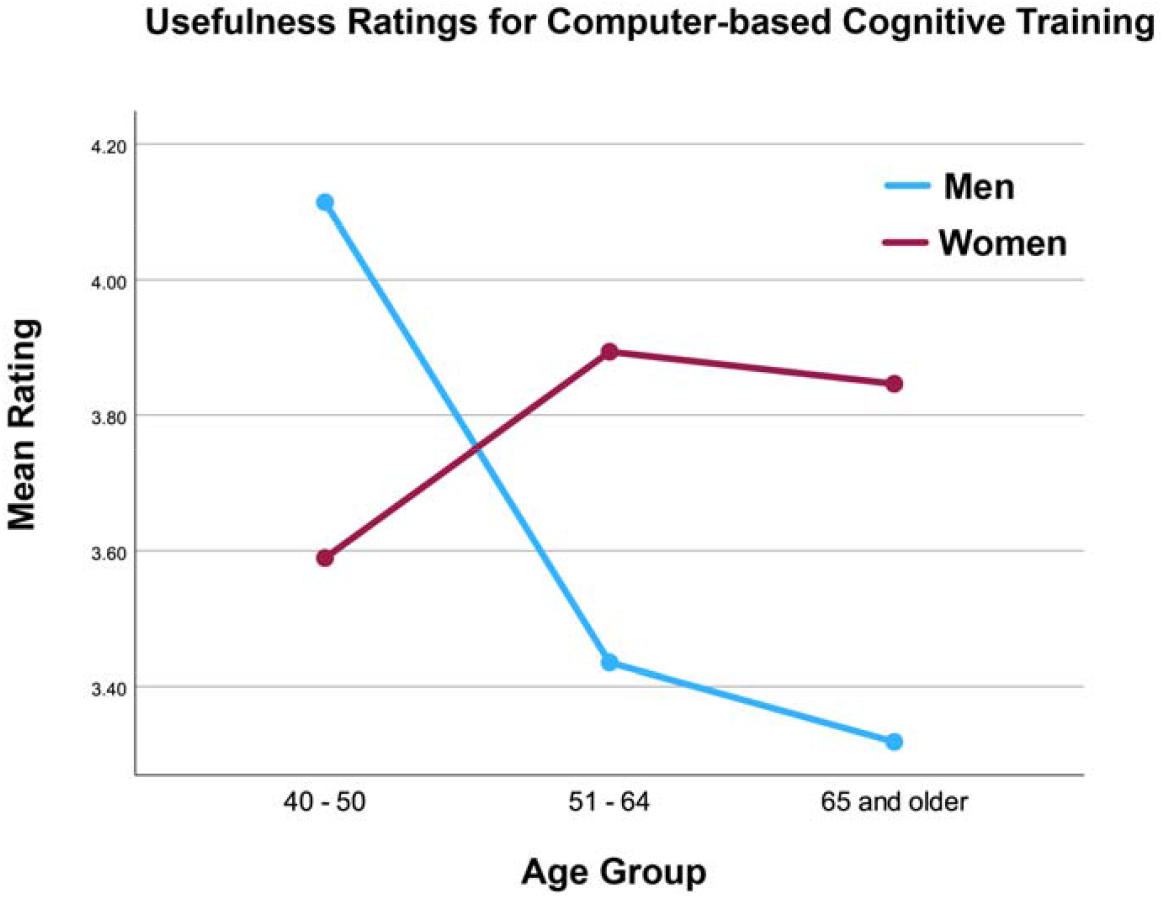
Interaction Between Age and Gender in Ratings of the Usefulness of Computer-Based Cognitive Training

### 3.2 Most Important Single Focus

In a separate section of the questionnaire, participants were asked, “If you were to focus on just one aspect of brain health, which would it be?” Across all respondents, aerobic exercise (*n* = 202) and mentally stimulating activities (*n* = 208) were the most frequently identified as the single most important activities for brain health. Other but less frequently rated activities included resistance training (*n* = 75), diet (*n* = 79), and sleep (*n* = 68).

### 3.3 Preferences for Service Delivery

Beyond activity-specific ratings, participants also reported their preferred modes of support for brain health interventions (Table 3). Just over half (52%) indicated a preference for working with someone who could assess their cognitive status, and more than one-third (36%) preferred a mobile app to assess memory and thinking. Fewer than 20% selected participation in an in-person group, while other preferred formats included online education (44%) and in-person education (29%).

**Table 3.** Preferences For Service Delivery.

|  |  |  |
| --- | --- | --- |
| Work with a provider who can give me a test | 393 | 51.6 |
| Online educational workshops or classes | 335 | 44.0 |
| App on my phone to test my memory and thinking | 277 | 36.4 |
| Work with a provider to develop a brain health plan | 226 | 29.7 |
| In person educational workshops or classes | 224 | 29.4 |
| App on my phone to track brain health activities | 207 | 27.2 |
| Reading a regular newsletter | 170 | 22.3 |
| Podcasts or video online | 157 | 20.6 |
| Working in a group face to face | 136 | 17.9 |
| Being able to text someone to get a quick answer | 128 | 16.9 |
| Online message board | 88 | 11.6 |
| E-mail consults with providers | 81 | 10.6 |
| Working in a group online | 60 | 10.5 |

## 4. Discussion

In this survey of middle-aged and older adults, we found that most participants endorsed a broad set of lifestyle behaviors as potentially helpful for maintaining or improving brain health. Activities tied to general well-being—such as good sleep, stress management, mentally stimulating activities, and social engagement—received consistently high ratings across age groups. In contrast, activities that require more effort, specialized skill, or sustained motivation, such as strength training, meditation, language learning, or computer-based cognitive training, were viewed as less helpful. Participants also showed strong interest in receiving some form of structured guidance, with more than half reporting a preference for cognitive status assessments and many indicating interest in online education or mobile app–based tools. Together, these findings suggest that adults who are concerned about cognitive aging are not only aware of at least some of the lifestyle factors that matter for brain health but are also open to receiving support in multiple formats.

Our results extend previous work showing that older adults have mixed yet generally optimistic beliefs about modifiable risk factors. Prior research indicates that adults commonly recognize mental stimulation, social engagement, and physical activity as protective [5, 7], while showing lower awareness of vascular risk factors, hearing loss, and mood-related contributors [8-10]. The high ratings of sleep and stress management in the present study may signal increasing public awareness of these factors, possibly driven by broader media attention and recent public health messaging. Likewise, participants’ strong ratings of mentally stimulating activities are consistent with reviews showing that such activities are widely endorsed as beneficial even when people do not always engage in them regularly [6]. The pattern of higher endorsement of multiple activities among younger men and older women mirrors gender- and age-linked differences in preventive health behavior reported elsewhere [23, 24], suggesting that interventions messages may need to be tailored to demographic subgroups.

The substantial interest in receiving cognitive assessments and online or app-based educational tools points to opportunities for scalable intervention delivery. Participants were less enthusiastic about group-based programs, especially in-person groups, echoing earlier work showing that older adults’ preferences for lifestyle programs vary widely and often reflect concerns about time, privacy, and convenience [11, 12]. The preference patterns observed here align with recommendations from public health entities, including the WHO and National Academies guidelines, emphasizing flexible, multicomponent approaches that integrate education, behavioral strategies, and risk-factor monitoring [15, 16]. Our findings also underscore the need for interventions that meet adults where they are—combining brief assessments, structured planning tools, and sustainable behavior-change supports rather than relying on single-component programs.

The prominence of aerobic exercise and mentally stimulating activities as the “one most important” activity for brain health is notable. These choices may reflect long-standing public messages about exercise and cognitive engagement, as well as their intuitive appeal. Yet the discrepancy between high ratings for activities like sleep or stress management and their lower selection as a top priority suggests that people may differentiate between what they believe is helpful and what they are most willing or able to focus on. This highlights the importance of helping adults set realistic priorities within a multicomponent lifestyle framework. An approach aligned with multidomain trials such as FINGER [3] and POINTER [25] and with public health models emphasizing small, cumulative behavior changes over time may be a reasonable strategy that can be tested to assess effectiveness.

This study has several limitations. Although the sample size was large, it was drawn from an online survey panel and was highly educated and mostly White, which may limit generalizability. Participants who volunteer for surveys on aging or health topics may be more motivated, more health literate, or more concerned about brain health than the general population. Ratings of activity usefulness were based on perceived rather than actual knowledge, and the concept of “usefulness” may vary across respondents. In addition, because the design was cross-sectional, we cannot infer how preferences or beliefs change with age or experience, nor can we determine whether expressed preferences translate into actual participation in brain health programs. Finally, although we examined age and gender differences, other sociodemographic factors—such as socioeconomic status, race/ethnicity, or caregiving experience—may also influence preferences and deserve further study.

Despite these limitations, this study provides potentially helpful information of what middle-aged and older adults know about brain health, what they prioritize, and how they would prefer to receive support. By linking knowledge, current behaviors, and intervention preferences, our results can inform the design of brain health programs that are both evidence-based and aligned with the needs and expectations of the communities they intend to serve. Future work should build on these findings by developing and testing multicomponent interventions that incorporate cognitive assessments, personalized planning, and flexible digital components, while ensuring accessibility for adults with varying levels of education, digital literacy, and health resources.

The preference for cognitive status assessments, reported by more than half of participants, may reflect a desire to reduce uncertainty or track changes in cognitive function. The absence of a clear consensus on preferred delivery formats underscores the importance of offering multiple, flexible options that account for individual differences. Intervention strategies that incorporate professional assessments, technology-based tools, and educational programs in both online and in-person formats may enhance adoption of brain-healthy behaviors. Future efforts should build on these insights by tailoring multicomponent, lifestyle-based brain health interventions to align with individual preferences and demographic patterns, which may improve engagement and long-term adherence.

## Author Contributions

Conceptualization, R.L.O.; methodology, R.L.O.; validation, R.L.O., J.C. and G.C.; formal analysis, R.L.O.; investigation, R.L.O.; data curation, R.L.O.; writing—original draft preparation, R.L.O.; writing—review and editing, G.C., S.A., S.W., J.C.; visualization, R.L.O., G.C.; All authors have read and agreed to the published version of the manuscript.

## Funding

This research received no external funding.

Institutional Review Board Statement: The study was conducted in accordance with the Declaration of Helsinki, and approved by the Institutional Review Board of Nova Southeastern University (protocol number NSU-2024-10, approval date January 9, 2024).

## Informed Consent Statement

Informed consent was obtained from all subjects involved in the study. Participants were required to review an online consent form detailing the purpose of the study and asked to provide their consent for participation after its review before responding to the survey.

## Data Availability Statement

The dataset contains individual-level survey responses from a small sample of older adults. Because the informed consent did not permit public data sharing and the IRB approval requires protection of participant confidentiality, the dataset cannot be made publicly available. Public posting could allow deductive re-identification even after de-identification procedures. Researchers may request access to a minimally de-identified dataset from the authors subject to a data use agreement

## Acknowledgment

During the preparation of this manuscript/study, the author(s) used ChatGPT 5.0 and 5.1 for the purposes of developing initial drafts of some sections. All portions of the manuscript were reviewed and completely rewritten. The authors have reviewed and edited the output and take full responsibility for the content of this publication.

## Disclaimer/Publisher’s Note

The statements, opinions and data contained in all publications are solely those of the individual author(s) and contributor(s) and not of MDPI and/or the editor(s). MDPI and/or the editor(s) disclaim responsibility for any injury to people or property resulting from any ideas, methods, instructions or products referred to in the content.

